# Maternal adiposity and inflammatory immune trajectories during pregnancy in women with HIV

**DOI:** 10.64898/2026.05.04.26352123

**Authors:** Jessica More, Elton Mukonda, Shameem Jaumdally, Hlengiwe P. Madlala, Clive Gray, Landon Myer, Marie-Louise Newell, Thokozile R. Malaba

## Abstract

Pregnancy requires tightly regulated immune adaptation, which may be altered by maternal adiposity and HIV infection. In high HIV prevalence settings with rising obesity rates, overlapping metabolic and infectious inflammation may shape gestational immune trajectories. We examined patterns of C-reactive (CRP), serum amyloid A (SAA), and interferon-gamma inducible protein-10 (IP-10) among 527 pregnant women living with HIV in Cape Town, South Africa. Immune markers were measured at up to four antenatal visits. Maternal body mass index (BMI) was categorised as normal, overweight or obese. Antiretroviral therapy (ART) exposure was classified as initiation before conception or during pregnancy. Mixed-effects models assessed associations, adjusting for baseline CD4 cell count and viral load.

CRP concentrations remained elevated across pregnancy and were significantly higher among obese women independent of ART timing. In contrast, IP-10 concentrations were strongly associated with ART timing, with higher levels observed prior to ART initiation and declining thereafter; no independent association with BMI was observed. SAA concentrations were modestly lower among obese women and among those on preconception ART. These divergent patterns persisted after adjustment for HIV disease severity.

These findings suggest that metabolic and HIV-related inflammatory pathways operate in parallel during pregnancy, with adiposity predominantly influencing acute-phase responses and HIV-related immune activation shaping interferon-associated chemokines trajectories.

## 1. Introduction

Maternal obesity and HIV infection are highly prevalent among pregnant women in South Africa and represent overlapping inflammatory exposures during gestation. Over 40% of pregnant South African women are classified as obese at the first antenatal care visit (ANC) (Madlala et al., 2021). In the Prematurity Immunology in HIV-infected Mothers and their Infants Study (PIMS) cohort, we previously demonstrated that elevated maternal BMI and gestational weight gain are significantly associated with adverse birth outcomes, underscoring the importance of characterising the underlying inflammatory milieu (Madlala et al., 2020). Obesity in pregnancy is characterised by chronic low-grade systemic inflammation and activation of placental inflammatory pathways, including macrophages infiltration and altered vascular signalling (Aye et al., 2014; Denison et al., 2010). Independently, maternal HIV infection is associated with persistent immune activation, even among women receiving antiretroviral therapy (ART)(Akoto et al., 2021; Vyas et al., 2021).

In pregnancy, immune adaptation requires tightly regulated shifts in inflammatory signalling to support implantation, placental vascularisation, and fetal development (Yockey & Iwasaki, 2018). Both metabolic dysregulation and chronic viral infection may perturb these processes. In the PIMS study, initiation of ART prior to conception was associated with approximately a two-fold increased risk of maternal vascular malperfusion (MVM) in the placenta, and MVM was independently association with adverse birth outcomes (Ikumi et al., 2021). These findings suggest that ART timing and maternal immune status may influence placental vascular pathology through inflammatory pathways.

Circulating inflammatory mediators provide insight into systemic immune activation during pregnancy. Interferon-gamma-inducible protein-10 (IP-10; CXCL10) reflects interferon-driven immune activation and T cell recruitment and is elevated in HIV infection, correlating with immune activation and CD4+ T cell depletion (Lei et al., 2019; Noel et al., 2014). During pregnancy, interferon signalling plays a central role in angiogenesis and maternal-fetal immune interactions (Yockey & Iwasaki, 2018), making IP-10 a biologically relevant proxy for HIV-associated immune perturbation that often remains elevated despite suppressive ART (Schnittman et al., 2021).

C-reactive protein (CRP) and serum amyloid A (SAA) are acute-phase reactants that index systemic inflammatory activation. CRP is strongly associated with adiposity and chronic metabolic inflammation (Fain, 2006; Reilly & Saltiel, 2017), and rises physiologically during late gestation and parturition (Aghaeepour et al., 2017; Mor & Cardenas, 2010). Among pregnant women living with HIV (WLWH), CRP levels have been associated with systemic immune activation and may vary according to ART regimen (Hindle et al., 2023; Vyas et al., 2021). SAA increasingly recognised as an adipose-associated inflammatory mediator (Yang et al., 2006), has been proposed as a danger associated molecular pattern in pregnancy (Lin et al., 2022). However, its longitudinal behaviour in WLWH and its relationship to maternal adiposity and ART timing remain poorly defined within the gestational period (Hrolfsdottir et al., 2016).

Together, IP-10, CRP, and SAA capture complementary inflammatory axes relevant to HIV-associated immune activation and obesity-related metabolic inflammation. While these markers do not represent comprehensive immune phenotyping, they allow evaluation of how maternal adiposity and ART timing are associated with systemic inflammatory trajectories across gestation in a high HIV- and obesity-prevalence setting.

We therefore examined longitudinal patterns of IP-10, CRP, and SAA according to maternal BMI category and ART initiation timing in a prospective cohort of pregnant WLWH in Cape Town, South Africa.

## 2. Materials and Methods

### 2.1 Study design and participants

This was a longitudinal analysis within the Prematurity Immunology in HIV-infected Mothers and their infants Study (PIMS), a prospective cohort conducted in Cape Town, South Africa. Pregnant women living with HIV (WLWH) were enrolled at ≤24 weeks’ gestation at their first antenatal care visit and followed through pregnancy with repeated clinical and immunological assessments. Details of the PIMS study design have been published previously(Malaba et al., 2021).

Participants were recruited between April 2015 and October 2016 from a large public-sector primary care antenatal care facility serving a high HIV-prevalence community. Gestational age was confirmed by research ultrasound at enrolment. Women were eligible if they were 18 years or older and planning to remain in the study area for delivery

Ethical approval was obtained from the University of Cape Town Faculty of Health Sciences Human Research Ethics Committee and the University of Southampton Institutional Review Board. All participants provided written informed consent.

### 2.2 Data collection and measurements

Participants underwent intensive measurements across study visits throughout pregnancy by trained study staff. Maternal sociodemographic, obstetric, and clinical data were collected at enrolment using structured questionnaires and abstraction from routine antenatal records. Phlebotomy was conducted at enrolment (visit 1: <24 weeks’ gestation and pre-ART initiation for those initiating ART at first antenatal visit), visit 2 (for those initiating ART in pregnancy an additional study visits two weeks post-ART initiation), visit 3 (28 weeks’ gestation for all) and visit 4 (34 weeks’ gestation for all). Anthropometric measurements, including maternal weight and height, were obtained using standardised equipment: calibrated scale (Charder, Taichung City, Taiwan) accurate to within 0.5 kg and a stadiometer (Seca, Birmingham, UK) accurate to the nearest 0.1.

### 2.3 Laboratory procedures

CRP and SAA were selected as markers of acute-phase inflammatory activation, while IP-10 was selected as a marker of interferon-mediated immune activation relevant to HIV-associated immune perturbation during pregnancy.

Plasma concentrations of SAA, CRP, and IP-10 were measured using quantitative sandwich ELISA kits (SAA: Novus Biologicals, NBP2-68119; CRP and IP-10: R&D Systems Quantikine) according to manufacturer protocols. Reported assay sensitivities were 0.75 ng/mL (SAA), 0.022 ng/mL (CRP), and 4.46 pg/mL (IP-10). Based on preliminary optimisation experiments, CRP was assayed at 1:100 dilution, while SAA and IP-10 were analysed undiluted.

All longitudinal samples from each participant were analysed on the same plate to minimise inter-assay variability. Inter- and intra-assay reliability were assessed using duplicated samples and inter-plate controls, with coefficients of variation (CV) calculated for each analyte. Intra-assay CVs were consistently <10% across all analytes, confirming high technical reproducibility.

CRP concentrations are reported in mg/L for clinical interpretability (1mg/L = 1000 ng/mL)

### 2.4 Exposures and Outcomes

#### 2.4.1 Maternal Adiposity

BMI was calculated at enrolment using measured height and weight, and categorised based on World Health Organization (WHO) classifications (WHO, 2000):

- Normal (<25.0 kg/m^2^)
- Overweight (25.0-29.9 kg/m^2^)
- Obese (≥30 kg/m^2^)

#### 2.4.2 ART exposure

ART exposure was categorised by timing of ART initiation:

- Preconception ART: initiated before pregnancy
- Pregnancy ART: initiated during current pregnancy

#### 2.4.3 Outcomes

The primary outcomes were longitudinal plasma concentrations of the three immune markers. These included CRP reported in mg/L with a predefined threshold for elevation at ≥10 mg/L (as a marker for high systemic risk); SAA reported in ng/mL, and IP-10 reported in pg/mL.

### 2.5 Statistical analysis

Descriptive statistics were used to summarise participant characteristics and baseline immune marker concentrations. Continuous variables are presented as median and interquartile ranges (IQR), and categorical variables as proportions. Normality was assessed using the Shapiro-Wilks test. Differences in immune marker concentrations across BMI categories at individual visits were evaluated using the Kruskal-Wallis test, with post-hoc Dunn’s test conducted where significant differences were observed.

To mitigate potential bias from incomplete data, multiple imputation by chained equations (MICE) was performed on predictor variables using 150 imputed datasets. The method of predictive mean matching (PMM) was used for continuous exposures and logistic regression for categorical variables. Biomarker variables were log-transformed to achieve normality and were not imputed. Missingness patterns were assessed prior to imputation.

Longitudinal associations between maternal adiposity, ART timing and immune marker concentrations were assessed using generalised linear mixed-effects models (GLMM) with random intercepts. Separate models were constructed for CRP, SAA, and IP-10, treating immune marker levels as time dependent outcomes. To address potential confounding, adjusted models included BMI category, ART timing, visit (gestational age time point), baseline CD4 cell count, baseline viral load and maternal age. These covariates were specified *a priori* to distinguish adiposity-driven responses from HIV-related inflammatory trajectories.

Results are presented as beta (β) coefficients with 95% confidence intervals (CI). Statistical significance was defined as p < 0.05. Analyses were performed using Stata version 17.0 (StataCorp, College Station, TX, USA) and R Studio.

## 3 Results

### 3.1 Participant characteristics

A total of 527 pregnant WLWH were included in this analysis. Median gestational age at enrolment was 14 weeks (IQR 11–18). Overall, 78% of participants were overweight or obese at baseline.

Obese participants were older (median 31 years) and more likely to be multigravida compared with women of normal BMI. At baseline, 66% of the cohort had suppressed viral load (<50 copies/mL), and median CD4 cell count was 433 cells/µL. ART timing was evenly distributed (53% preconception, 47% pregnancy initiation) and did not differ across BMI categories (Table 1).

**Table 1.** Baseline demographic, obstetric, and HIV-related characteristics of pregnant women living with HIV (n = 527), stratified by BMI category.

|  | **Total**<br>**N=527** | **Normal**<br>**N=115** | **Overweight**<br>**N=143** | **Obese**<br>**N=269** |
| --- | --- | --- | --- | --- |
| **Maternal Characteristics** |  |  |  |  |
| Age, years |  |  |  |  |
| ≤24 | 88 (17) | 28 (24) | 22 (15) | 38 (14) |
| 25-29 | 151 (29) | 37 (32) | 45 (31) | 69 (26) |
| ≥30 | 288 (55) | 50 (43) | 76 (53) | 162 (60) |
| Median (IQR) | 30 (26.0-34.0) | 28 (25.0-33.0) | 30 (26.0-34.0) | 31 (27.0-35.0) |
| **Obstetric Characteristics** |  |  |  |  |
| Gravidity |  |  |  |  |
| 1 | 87 (17) | 27 (24) | 25 (18) | 35 (13) |
| 2 | 178 (34) | 43 (38) | 51 (36) | 84 (32) |
| ≥3 | 255 (49) | 43 (38) | 65 (46) | 147 (55) |
| Missing | 7 (1) | 2 (2) | 2 (1) | 3 (1) |
| Parity |  |  |  |  |
| 0 | 124 (24) | 37 (33) | 35 (25) | 52 (20) |
| 1 | 214 (41) | 49 (43) | 52 (37) | 113 (43) |
| ≥2 | 124 (24) | 37 (33) | 35 (25) | 52 (20) |
| Missing | 8 (2) | 2 (2) | 2 (2) | 4 (2) |
| Gestational age (weeks), median (IQR) | 14 (11-18) | 14.5 (11-18) | 15 (11-18) | 14 (10-17) |
| Haemoglobin (g/dl), median (IQR) | 11.1 (10.2-12.0) | 10.3 (9.8-11.6) | 11.0 (10.1-11.8) | 11.5 (10.6-12.1) |
| Missing | 63 (12) | 13 (11) | 18 (13) | 32 (12) |
| **HIV** |  |  |  |  |
| ART status (timing of initiation) |  |  |  |  |
| Before pregnancy | 278 (53) | 60 (52) | 76 (53) | 142 (53) |
| During current pregnancy | 249 (47) | 55 (48) | 67 (47) | 127 (47) |
| ART regimen, self-report |  |  |  |  |
| TDF-3TC-EFV | 493 (94) | 110 (95) | 132 (92) | 251 (93) |
| Other NNRTI-based regimen | 24 (4) | 3 (3) | 7 (5) | 14 (5) |
| PI-based regimen | 10 (2) | 2 (2) | 4 (3) | 4 (1) |
| CD4 cell count, cells/μL* |  |  |  |  |
| ≤ 200 | 45 (9) | 12 (10) | 16 (11) | 17 (6) |
| 201-350 | 97 (18) | 23 (20) | 24 (17) | 50 (19) |
| 351-500 | 110 (21) | 27 (24) | 34 (24) | 49 (18) |
| >500 | 167 (32) | 38 (33) | 41 (29) | 88 (33) |
| Missing | 108 (21) | 15 (13) | 28 (20) | 65 (24) |
| Median (IQR) | 433 (298-600) | 435 (294-568) | 411 (302-589) | 467 (303-626) |
| VL, copies/mL* |  |  |  |  |
| <50 | 346 (66) | 73 (63) | 88 (62) | 185 (69) |
| 50-1000 | 66 (13) | 16 (14) | 22 (15) | 28 (10) |
| >1000 | 59 (11) | 17 (15) | 15 (11) | 27 (10) |
| Missing | 56 (11) | 9 (8) | 18 (13) | 29 (11) |
| Median (IQR) | 20 (20-67) | 20 (20-150) | 20 (20-88) | 20 (20-31) |
| Previous TB, n (%) |  |  |  |  |
| Yes | 88 (19) | 20 (19) | 25 (20) | 43 (18) |
| No | 381 (81) | 86 (81) | 100 (80) | 195 (82) |
| Missing | 58 (11) | 9 (8) | 18 (13) | 31 (12) |

Data are presented as n (%) or median (IQR). CD4 cell count and viral load values were abstracted from routine clinical records and represent measurements closest to the first antenatal visit.
Abbreviations: ART, antiretroviral therapy; BMI, body mass index; IQR, interquartile range; VL, viral load; TB, tuberculosis.

Imputation diagnostics demonstrated stability of model estimates and comparable distributions between observed and imputed values (Supplementary Figure S1).

### 3.2 Longitudinal inflammatory marker profiles by BMI

CRP concentrations remained elevated throughout pregnancy, with higher levels observed among obese women across visits. SAA concentrations demonstrated less pronounced variation over gestation, while IP-10 concentrations showed temporal changes primarily among women initiating ART during pregnancy.

Across BMI categories, median CRP concentrations were consistently higher among obese participants compared with women of normal BMI (Table 2). These longitudinal patterns are illustrated in Figure 1.

**Figure 1.**
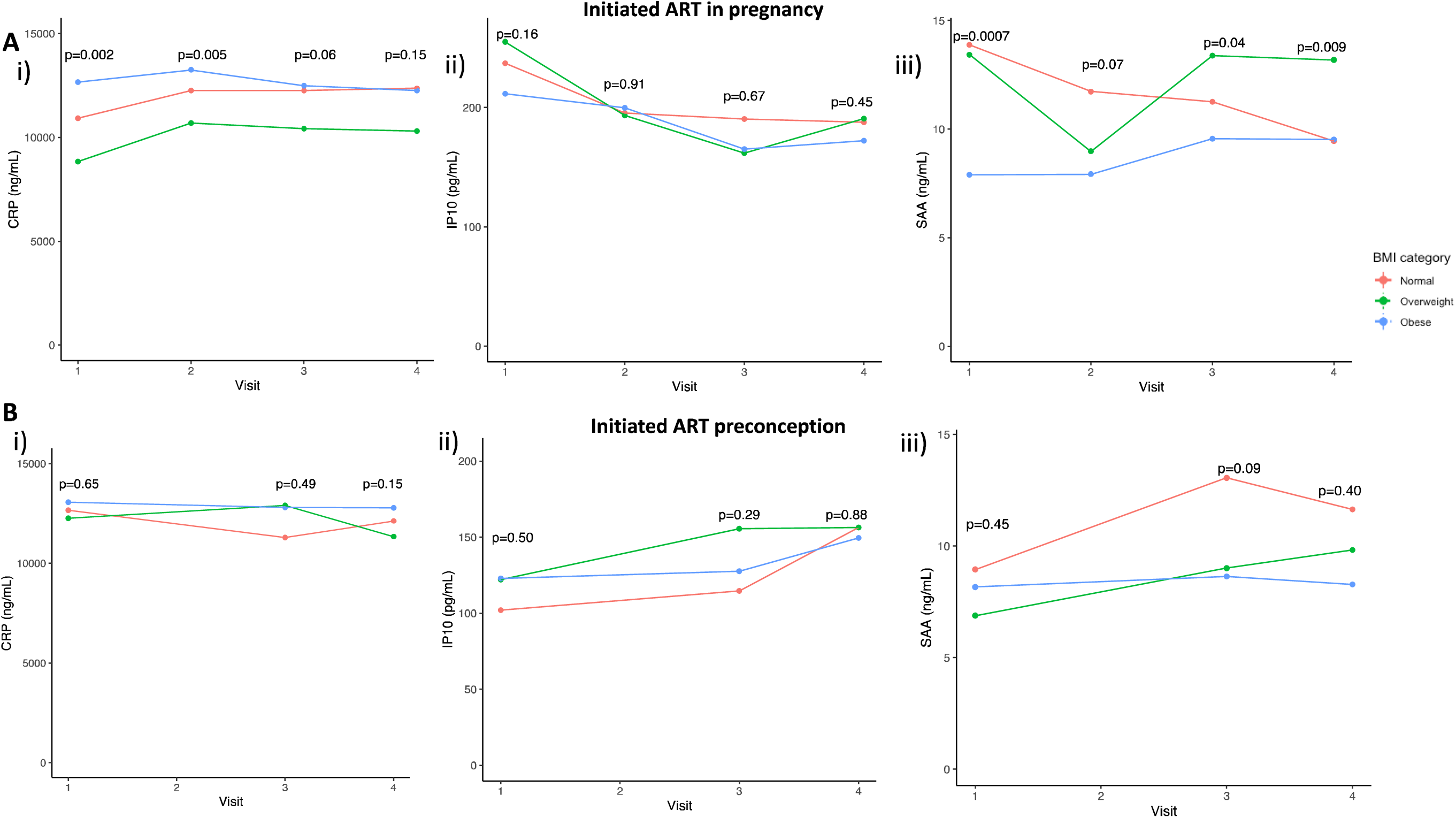
Longitudinal plasma concentrations of inflammatory markers by BMI category and ART timing. Median concentrations of (A) C-reactive protein (CRP), (B) interferon-γ-inducible protein-10 (IP-10), and (C) serum amyloid A (SAA) across pregnancy, stratified by BMI category (normal, overweight, obese). Panels are shown separately for women initiating ART during pregnancy and those on preconception ART. Study visits include enrolment (Visit 1), approximately 2 weeks post-ART initiation for those initiating during pregnancy (Visit 2), ∼28 weeks’ gestation (Visit 3), and ∼34 weeks’ gestation (Visit 4). CRP is reported in mg/L; SAA in ng/mL; IP-10 in pg/mL. Abbreviations: ART, antiretroviral therapy; BMI, body mass index.

**Table 2.** Median (IQR) plasma concentrations of inflammatory markers across pregnancy by BMI category.

| Immune marker by Visit | Total (N) | Normal (N) | Overweight (N) | Obese (N) | P*** |
| --- | --- | --- | --- | --- | --- |
| **Visit 1** | **527** | **115** | **143** | **269** |  |
| **Gestational age\*, Median (IQR)** | 14 (11 - 18) | 14.5 (11 - 18) | 15 (11.25 - 18) | 14 (10 - 17) | 0.20 |
| **CRP (mg/L)** | 12.41 (75.47 - 15.16) | 11.38 (5.89 - 14.65) | 10.76 (6.38 - 14.98) | 12.78 (9.19 - 16.99) | **0.05** |
| **IP-10 (pg/mL)** | 160.76 (105.46 - 259.36) | 163.35 (97.37 - 267.39) | 161.59 (102.29 - 258.35) | 153.39 (110.92 - 258.91) | 0.76 |
| **SAA (ng/mL)** | 8.93 (4.80 - 16.33) | 9.74 (5.32 - 20.29) | 9.99 (4.68 - 17.48) | 8.08 (4.82 - 13.84) | 0.10 |
| **Visit 2\*\*** | **223** | **46** | **62** | **115** |  |
| **Gestational age\*, Median (IQR)** | 17 (14 - 21) | 17 (14 - 21) | 18 (15 - 21) | 16 (13 - 20) | 0.18 |
| **CRP (mg/L)** | 12.78 (8.02 - 16.5) | 12.25 (8.02 - 14.64) | 10.69 (6.72 - 15.83) | 13.43 (11.40 - 17.07) | **0.01** |
| **IP-10 (pg/mL)** | 197.84 (139.13 - 274.96) | 195.35 (127.56 - 266.96) | 193.41 (139.56 - 296.52) | 199.72 (141.94 - 274.96) | 0.89 |
| **SAA (ng/mL)** | 8.78 (5.32 - 15.12) | 11.73 (6.75 - 18.70) | 8.98 (4.43 - 17.81) | 7.93 (5.33 - 13.61) | 0.34 |
| **Visit 3** | **440** | **63** | **128** | **249** |  |
| **Gestational age\*, Median (IQR)** | 28 (27 - 28) | 27 (27 - 28) | 27 (27 - 28) | 27 (27 - 28) | 0.92 |
| **CRP (mg/L)** | 12.31 (8.07 - 15.28) | 12.25 (8.12 - 14.98) | 11.58 (7.34 - 15.16) | 12.75 (8.57 - 15.60) | 0.22 |
| **IP-10 (pg/mL)** | 148.71 (105.49 - 214.39) | 145.01 (100.48 - 234.91) | 157.82 (114.20 - 216.53) | 143.84 (103.95 - 208.70) | 0.40 |
| **SAA (ng/mL)** | 10.02 (5.12 - 16.69) | 11.71 (6.95 - 19.07) | 11.05 (5.80 - 19.05) | 9.21 (4.72 - 15.08) | 0.12 |
| **Visit 4** | **415** | **49** | **116** | **250** |  |
| **Gestational age\*, Median (IQR)** | 34 (33 - 34) | 34 (33 - 34) | 34 (33 - 34) | 33 (33 - 34) | 0.89 |
| **CRP (mg/L)** | 12.55 (7.81 - 14.28) | 12.31 (9.20 - 14.51) | 10.70 (5.56 - 12.90) | 12.66 (8.56 - 14.98) | **0.03** |
| **IP-10 (pg/mL)** | 164.51 (127.43 - 229.90) | 179.98 (130.31 - 223.21) | 172.04 (130.88 - 250.99) | 158.76 (123.74 - 217.23) | 0.51 |
| **SAA (ng/mL)** | 9.97 (5.78 - 16.70) | 10.61 (7.44 - 16.58) | 11.07 (6.35 - 18.50) | 8.55 (5.17 - 16.05) | 0.33 |

### 3.3 Acute-phase markers by obesity class

Stratification by obesity class (I, II, and III) demonstrated elevated CRP concentrations across all classes compared with normal BMI. However, no clear dose-response relationship was observed between increasing obesity severity and inflammatory marker concentrations.

SAA concentrations showed modest variation across obesity classes. These stratified patterns are presented in Figure 2.

**Figure 2.**
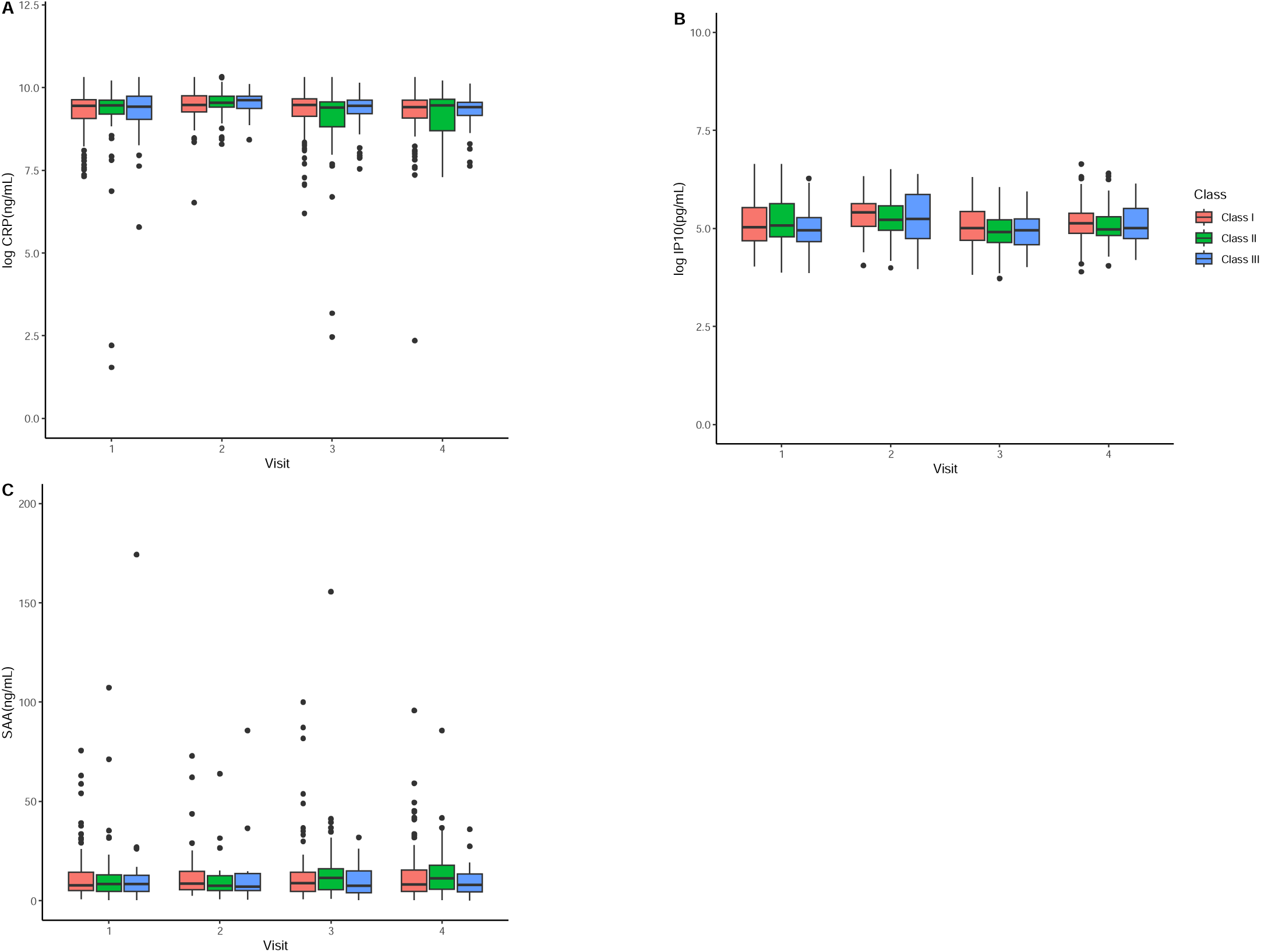
Longitudinal inflammatory marker concentrations by obesity class. Median concentrations of (A) CRP, (B) IP-10, and (C) SAA across pregnancy among women classified as obesity class I, II, or III. Visits correspond to enrolment (Visit 1), approximately 2 weeks post-ART initiation for women initiating ART during pregnancy (Visit 2), ∼28 weeks’ gestation (Visit 3), and ∼34 weeks’ gestation (Visit 4). CRP is reported in mg/L; SAA in ng/mL; IP-10 in pg/mL. Abbreviations: BMI, body mass index; ART, antiretroviral therapy.

### 3.4 Interferon-associated chemokine trajectories by ART timing

IP-10 concentrations differed according to ART timing. At baseline, women initiating ART during pregnancy had higher IP-10 concentrations compared with women on preconception ART. Following ART initiation, IP-10 concentrations declined in the pregnancy-initiation group, whereas levels among preconception ART users remained relatively stable.

CRP concentrations remained elevated across visits regardless of ART timing, particularly among obese participants. These patterns are shown in Table 3 and Figure 3.

**Figure 3.**
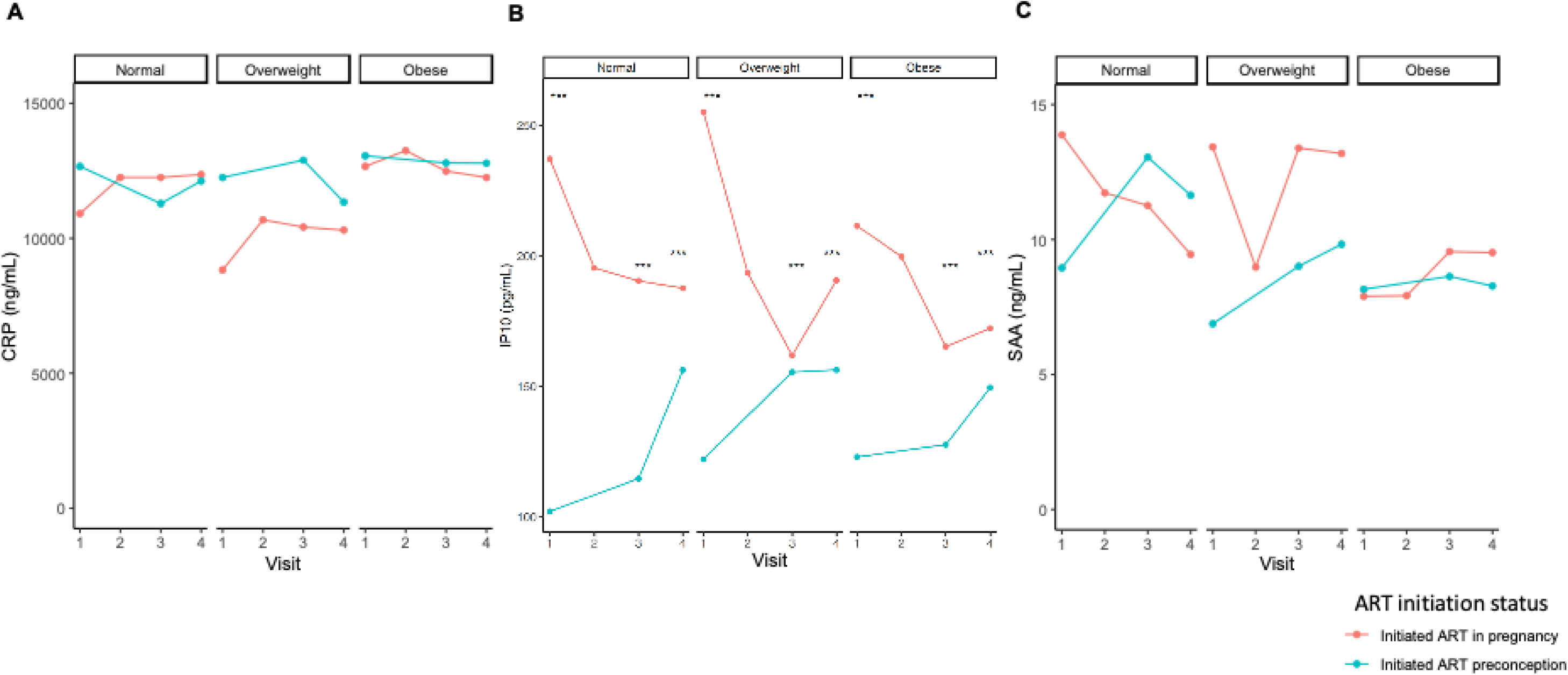
Inflammatory marker trajectories by ART timing stratified by BMI category. Median concentrations of (A) CRP, (B) IP-10, and (C) SAA across pregnancy among women initiating ART during pregnancy and those on preconception ART, stratified by BMI category. Visits include enrolment (Visit 1), approximately 2 weeks post-ART initiation (Visit 2; pregnancy initiation group only), ∼28 weeks’ gestation (Visit 3), and ∼34 weeks’ gestation (Visit 4). CRP is reported in mg/L; SAA in ng/mL; IP-10 in pg/mL. Abbreviations: ART, antiretroviral therapy; BMI, body mass index.

**Table 3.** Median (IQR) plasma concentrations of inflammatory markers across pregnancy by ART initiation timing.

| Immune marker by Visit | Total (N) | Initiated in current pregnancy (N) | Initiated preconception (N) | P |
| --- | --- | --- | --- | --- |
| **Visit 1** | **527** | **278** | **249** |  |
| **Gestational age (GA), weeks**<br>**Median (IQR)** | 14 (11 - 18) | 14 (11 - 18) | 14 (11 - 17) | 0.29 |
| **CRP (mg/L)** | 12.41 (7.54 - 15.16) | 11.65 (7.11 - 14.99) | 12.84 (7.72 - 15.17) | 0.18 |
| **IP-10 (pg/mL)** | 160.76 (146.02 - 259.35) | 226.97 (147.72 - 353.29) | 121.05 (86.72 - 171.33) | **<0.001** |
| **SAA (ng/mL)** | 8.93 (4.79 - 16.33) | 9.29 (4.84 - 17.38) | 8.16 (4.67 - 15.37) | **0.04** |
| **Visit 2\*** | **223** | **223** |  |  |
| **Gestational age (GA), weeks**<br>**Median (IQR)** | 17 (14 - 21) | 17(14 - 21) |  |  |
| **CRP (mg/L)** | 12.78 (8.02 - 16.59) | 12.78 (8.02 - 16.59) |  |  |
| **IP-10 (pg/mL)** | 197.84 (139.13 - 274.96) | 197.84 (139.13 - 274.96) |  |  |
| **SAA (ng/mL)** | 8.78 (5.32 - 15.12) | 8.78 (5.32 - 15.12) |  |  |
| **Visit 3** | **440** | **232** | **208** |  |
| **Gestational age (GA), weeks**<br>**Median (IQR)** | 27 (27 - 28) | 27 (27 - 28) | 27 (27 - 28) | **0.004** |
| **CRP (mg/L)** | 12.34 (8.09 - 15.30) | 12.25 (7.98 - 15.20) | 12.78 (8.16 - 15.28) | 0.67 |
| **IP-10 (pg/mL)** | 148.65 (105.50 - 214.12) | 165.23 (122.14 - 247.65) | 135.56 (99.29 - 182.17) | **<0.001** |
| **SAA (ng/mL)** | 10.01 (5.10 - 16.66) | 10.76 (5.69 - 18.34) | 9.23 (4.87 - 16.21) | 0.27 |
| **Visit 4** | **415** | **217** | **198** |  |
| **Gestational age (GA), weeks**<br>**Median (IQR)** | 34 (33 - 34) | 34 (33 - 34) | 34 (33 - 34) | **0.003** |
| **CRP (mg/L)** | 12.25 (7.81 - 14.28) | 12.09 (7.48 - 14.25) | 12.35 (8.49 - 14.70) | 0.45 |
| **IP-10 (pg/mL)** | 164.51 (127.43 - 229.90) | 179.88 (133.71 - 251.92) | 150.61 (116.16 - 201.20) | **0.004** |
| **SAA (ng/mL)** | 9.97 (5.78 - 16.70) | 10.24 (6.08 - 17.89) | 9.25 (5.36 - 15.66) | 0.55 |

### 3.5 Multivariable mixed-effects models

In adjusted mixed-effects models controlling for BMI category, ART timing, visit, baseline CD4 cell count, and viral load, obesity remained independently associated with CRP concentrations and inversely associated with SAA concentrations (Table 4). BMI category was not associated with IP-10 concentrations. In contrast, ART timing and viral load were independently associated with IP-10 concentrations but were not associated with CRP or SAA concentrations

**Table 4.** Adjusted mixed-effects models examining associations between BMI category, ART timing, visit, and inflammatory marker concentrations.

| *Predictors* | CRP (mg/L) |  |  | P-10 (pg/mL) |  |  | SAA (ng/mL) |  |  |
| --- | --- | --- | --- | --- | --- | --- | --- | --- | --- |
| | $\beta$ | CI | P | $\beta$ | CI | P | $\beta$ | CI | P |
| **BMI category** |  |  |  |  |  |  |  |  |  |
| Normal (ref) | - | - | - | - | - | - | - | - | - |
| Overweight | 0.26 | -0.58 – 1.10 | 0.55 | 3.23 | -16.27 - 22.73 | 0.75 | -1.47 | -4.34 - 1.40 | 0.32 |
| Obese | 1.28 | 0.32 – 2.2 | **<0.001** | -2.81 | -23.29 - 17.68 | 0.79 | -3.64 | -6.39 - -0.88 | **<0.001** |
| **ART status** |  |  |  |  |  |  |  |  |  |
| Initiated in pregnancy (ref) | - | - | - | - | - | - | - | - | - |
| Initiated preconception | 0.43 | -0.47 - 1.33 | 0.35 | -69.35 | -86.70 - -51.98 | **<0.001** | -2.26 | -4.43 - -0.09 | **0.04** |
| **Visit** |  |  |  |  |  |  |  |  |  |
| Visit 1 (ref) | - | - | - | - | - | - | - | - | - |
| Visit 2 | 1.52 | 0.91 - 2.13 | **<0.001** | -6.97 | -22.39 - 8.46 | 0.38 | -1.95 | -4.57 - 0.66 | 0.14 |
| Visit 3 | 0.23 | -0.25 - 0.71 | 0.35 | -31.45 | -43.48 - -19.41 | **<0.001** | -0.12 | -2.17 - 1.93 | 0.91 |
| Visit 4 | -0.21 | -0.70 - 0.28 | 0.40 | -12.62 | -24.97 - -0.27 | **0.05** | -0.47 | -2.57 - 1.62 | 0.66 |

These results indicate differential associations across inflammatory markers according to maternal adiposity and HIV-related clinical factors.

## 4. Discussion

### 4.1 Main findings

In this large cohort of pregnant WLWH attending ANC at a public sector facility in South Africa, we observed that proinflammatory immune marker profiles were shaped by both maternal BMI and timing of ART initiation. Specifically, CRP was consistently elevated above inflammatory thresholds across pregnancy, particularly among obese women, and remained high across gestation regardless of ART initiation timing. SAA concentrations were paradoxically lower among obese women especially in those on ART preconception. In contrast, IP-10 was primarily determined by HIV disease activity and ART initiation: in initiators levels were highest pre-ART at first ANC, declined following ART initiation. Among those already on ART levels rose moderately by third trimester. Adjusted analyses confirmed these trends with obesity the dominant driver of CRP and SAA levels, and HIV and ART status explaining IP-10 variation. These divergent trajectories suggest different underlying pathways of immune activation. These findings add to the growing evidence that immune dysregulation in pregnancy is shaped by the intersection of HIV, ART exposure, and maternal metabolic health.

### 4.2 Interpretation in context of existing literature

Pregnancy is characterised by tightly regulated inflammatory transitions that support placental development, vascular remodelling, and parturition (Aghaeepour et al., 2017; Mor & Cardenas, 2010). Both obesity and HIV infection are independently associated with chronic immune activation and may perturb these adaptive processes. Evidence from African maternal cohorts demonstrates that HIV exposure and ART use are associated with altered systemic immune mediator trajectories during pregnancy and postpartum (Akoto et al., 2021; Schnittman et al., 2021), supporting the broader concept that gestational immune adaptation in high-burden settings occurs within a background of chronic viral and metabolic stressors.

The consistent association between obesity and elevated CRP concentrations aligns with extensive evidence linking adiposity to chronic low-grade meta-inflammation (Fain, 2006; Reilly & Saltiel, 2017). Obesity has also been linked to placental inflammatory activation and increased placental macrophage accumulation (Challier et al., 2008), providing a biologically plausible connection between systemic meta-inflammation and altered placental function. In the PIMS cohort, higher maternal BMI was associated with adverse birth outcomes (Madlala et al., 2020), and ART initiation prior to conception was associated with increased risk of MVM (Ikumi et al., 2021). Although placental histopathology was not examined in the present analysis, sustained CRP elevation among obese WLWH may reflect systemic inflammatory states that plausibly interact with placental vascular processes. Given that MVM represents a defined placental lesion with standardised diagnostic criteria, future integration of systemic inflammatory markers with placental pathology could be anchored in established frameworks such as those defined by the Amsterdam Placental Workshop Group (Ernst, 2018; Khong et al., 2016).

In contrast, IP-10 trajectories were primarily associated with HIV disease markers and ART timing rather than BMI. IP-10 is a Th1-type interferon-inducible chemokine elevated in untreated HIV infection and correlated with immune activation and CD4+ T-cell depletion (Lei et al., 2019; Noel et al., 2014). Our observation that IP-10 concentrations were highest at enrolment among ART initiators and declined following ART initiation is consistent with longitudinal studies demonstrating ART-associated modulation of interferon-related mediators during pregnancy (Schnittman et al., 2021). Additional South African data indicate that systemic inflammatory mediators differ between WLWH and HIV-uninfected women (Sevenoaks et al., 2021), further reinforcing the sensitivity of interferon-associated pathways to HIV-related immune activation. The lack of association between BMI and IP-10 in our analysis suggests that interferon-driven immune activation may be more strongly influenced by viral and treatment-related factors than by metabolic status in this population.

SAA demonstrated an inverse association with obesity, which contrasts with findings from HIV-uninfected cohorts where SAA is typically elevated in obesity and with gestational weight gain (Hrolfsdottir et al., 2016; Yang et al., 2006). One potential explanation is altered acute-phase responsiveness in the setting of chronic HIV-associated immune activation, which persists despite suppressive ART and has been described as a state of sustained inflammatory and immunometabolic dysregulation in treated HIV infection (Teer et al., 2025). The biological significance of lower SAA concentrations in obese WLWH remains uncertain. Interpretation is further complicated by the high burden of co-endemic infectious exposures in sub-Saharan Africa, which may influence baseline inflammatory set-points during pregnancy.

Taken together, our findings indicate that maternal adiposity predominantly influences acute-phase inflammatory markers (CRP and SAA), whereas HIV disease activity and ART timing more strongly shape interferon-associated pathways reflected by IP-10. These divergent trajectories highlight that systemic inflammation during pregnancy in WLWH is pathway-specific rather than uniform, and that overlapping metabolic and virological exposures may influence distinct components of the maternal immune milieu.

### 4.3 Strengths and limitations

This study has several strengths. The large, well-characterised cohort with repeated sampling across four antenatal visits enabled detailed assessment of longitudinal inflammatory trajectories. Inclusion of women initiating ART during pregnancy and women on ART prior to conception allowed evaluation of treatment timing. Rigorous laboratory quality control, including high intra-assay reliability (CV <10%), minimised measurement variability.

Several limitations warrant consideration. The absence of an HIV-uninfected comparator group limits the ability to fully disentangle HIV-related from pregnancy-specific inflammatory changes. The biomarker panel was restricted to three circulating markers and does not represent comprehensive immune phenotyping. Although unsupervised clustering approaches were explored, limited separation was observed using a three-marker panel, reinforcing the interpretation that these markers reflect overlapping inflammatory processes rather than discrete immune phenotypes. BMI was measured at enrolment rather than pre-pregnancy, and longitudinal visit timing differed slightly between ART groups.

## 5. Conclusion

In pregnant women living with HIV, maternal adiposity and HIV-related clinical factors were associated with distinct systemic inflammatory trajectories across gestation. Maternal obesity was primarily associated with acute-phase inflammatory markers (CRP and SAA), whereas HIV disease markers and ART timing were more strongly associated with interferon-mediated immune activation reflected by IP-10. These findings indicate pathway-specific patterns of systemic inflammation in HIV-exposed pregnancy and underscore the importance of considering both metabolic and virological influences when examining immune regulation in high-burden settings. Future research should evaluate whether these divergent systemic inflammatory profiles are associated with specific placental pathologies or adverse birth outcomes.

## Data Availability

All data produced in the present study are available upon reasonable request to the authors.

## Acknowledgements

The authors are extremely grateful to all the women who participated in this study. We thank the staff at the Gugulethu Midwife Obstetric Unit for their support and extend our appreciation to the entire research team, including Colin Newell, Megan Mrubata, and Lee-Ann Stemmet, for their valuable contributions throughout the project.

## CRediT Author Statement

**MLN**: Conceptualisation, Funding acquisition, Writing (review & editing). **LM**: Conceptualisation, Project administration. **CG**: Conceptualisation, Methodology. **TRM**: Conceptualisation, Supervision, Writing (review & editing). **HPM**: Methodology (Maternal BMI component). JM: Formal analysis, Writing (original draft). **EM:** Formal analysis (statistical validation). **SJ**: Investigation (Laboratory assays), Data curation. All authors contributed to the interpretation of the findings, reviewed the manuscript, and approved the final version for submission.

## Ethics Approval

This study received ethical approval from the University of Cape Town Faculty of Health Sciences Human Research Ethics Committee (HREC; reference number: 734/2014) and the University of Southampton Institutional Review Board (reference number: 12542 PIMS). All study procedures were performed in accordance with the ethical standards of the institutional and/or national research committee and with the 1964 Helsinki Declaration and its later amendments.

