## Supplementary files for "Maternal adiposity and inflammatory immune trajectories during pregnancy in women with HIV"

**SUPPLEMENTARY DATA**

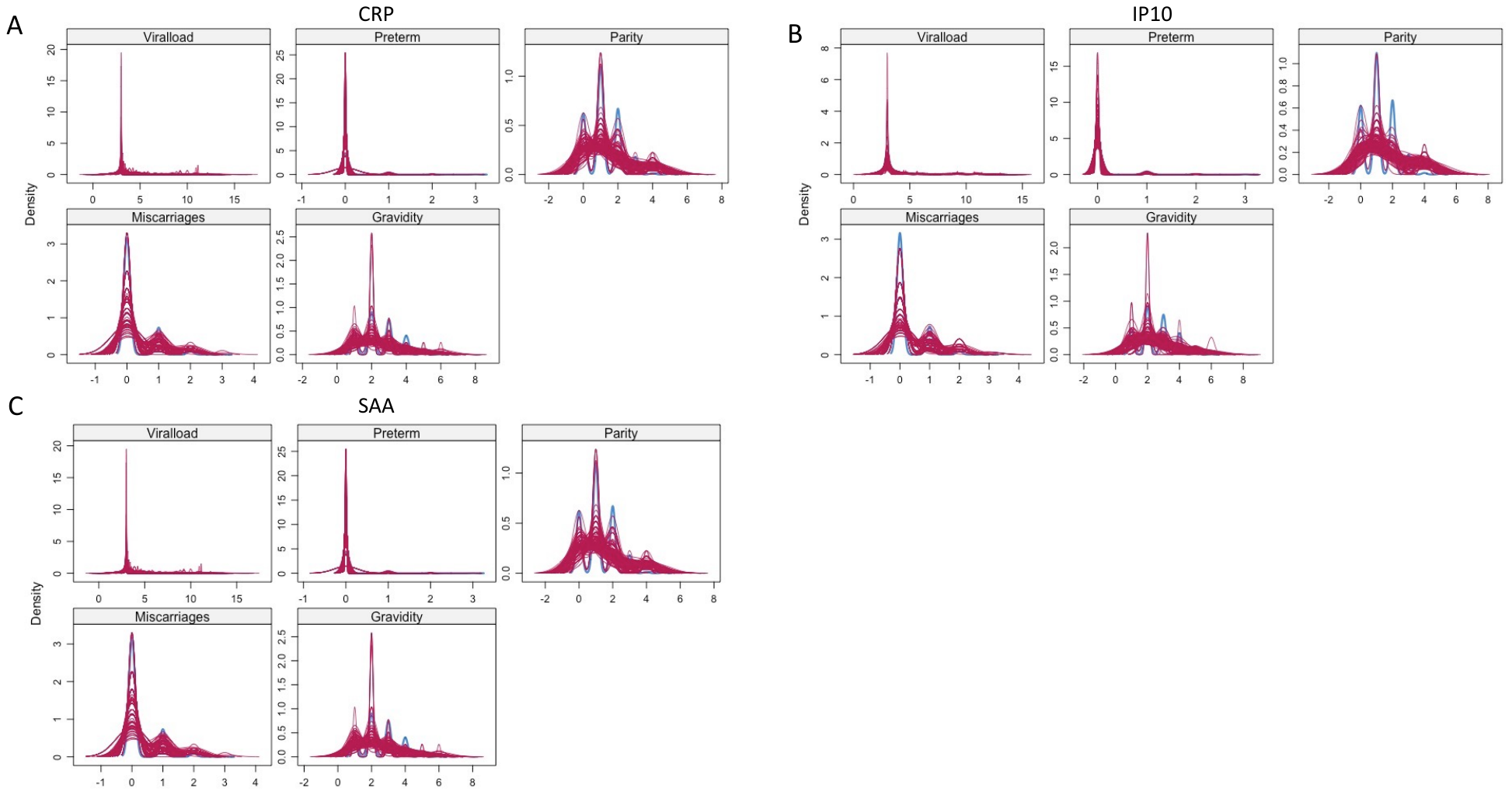

**Supplementary Figure S1.** Density plots showing the distribution of the observed and imputed data. The red (imputed data) and blue (observed data) for the biomarker outcome data similar densities in data with A) CRP, B) IP10 and C) SAA as the outcome. Abbreviations: CRP, C-reactive protein; SAA, Serum amyloid A; IP10, interferon-gamma-inducible protein 10.
